# Attitude towards gender inequitable norms and associated factors among male youths in Arba Minch town, Southern Ethiopia

**DOI:** 10.1101/2022.04.08.22273620

**Authors:** Barkot Samuel, Mulugeta Shegaze, Muluken Bafa Basa, Zeleke Gebru, Belay Boda, Mustefa Glagn

## Abstract

**Background:** Gender inequitable norms are common cultural traditions across the world, which characterized women as someone who deserves less basic life support and fundamental human rights than men. The unfairness of such practices has demolished women’s quality of life. Studies are limited to measuring some factors in Ethiopia. The information obtained from this study will help to take an evidence-based approach to reduce gender inequitable norms.

**Objective:** To assess attitude towards gender inequitable norms and associated factors among male youths from August 16^th^– 30^th^, 2020 at Arba Minch town, Southern Ethiopia.

**Methods:** A community-based cross-sectional study was conducted among 423 male youths. Gender-Equitable Men Scale was used to measure the attitudes towards gender inequitable norms. The study participants were selected using a simple random sampling technique by allocating a proportion to each kebeles. Both bivariable and multivariable logistic regression analyses were performed to identify associated factors. The level of statistical significance was set at a p-value of less than 0.05 in multivariable logistic regression.

**Results:** The overall prevalence of favorable attitude towards gender inequitable norms among male youths was 47.3% with 95% CI (42%, 52%). No formal education, lower household income, not participating in youth clubs, poor knowledge, and having a prior history of physical violence during childhood were factors significantly associated with a favorable attitude towards gender inequitable norms.

**Conclusion and recommendation:** Almost half of the participants had a favorable attitude towards gender inequitable norms. Proactive measures need to be taken to curb the problem by taking integrated interventions for youths like enrolling in school, enhancing participation in a youth club, prevent childhood physical violence, increasing household income, and creating awareness on gender inequitable norms to change traditional beliefs that undermine women.

## Introduction

Gender inequitable norms are socially constructed informal rules among a set of people and in most cultures and societies, they demean women and girls. They are context- and time-specific, changeable, learned through socialization processes, and govern people [1, 2]. Most victims of gender inequitable norms (GINs) are women and girls and its root cause is men’s or boys’ attitude towards gender inequitable norms [3].

Men’s and women’s roles and attitudes according to gender are classified as democratic and traditional. The traditional roles have given women and men unequal responsibilities, which encourage behaviors that place men and their sexual partners at risk of various negative health outcomes [4, 5]. In countries where gender-equitable norms are not ensured, all practiced gender norms have negative impacts like high maternal mortality and deaths among women than men, unintended pregnancy, STIs, HIV/AIDS, and barriers to the utilization of reproductive health services and teenage pregnancies [6, 7].

Few studies were conducted in different cultural settings to identify factors associated with an attitude towards gender inequitable norms, which included age, academic achievement, marital status, employment, and decision making [8, 9]. Though those studies list out the factors that can affect the woman related to gender inequitable norms, they didn’t mention what makes society more vulnerable to such attitude [10]. A clear understanding of factors associated with gender inequitable norms specifically by targeting male youth groups of the population is important to help in the development and implementation of evidence-based approaches and for various interventions to reduce gender inequitable norms. Thus, this study assessed attitude towards gender inequitable norms and associated factors among male youths in Arba Minch town using Gender Equitable Men Scale (GEMS).

## Materials and methods

### Setting and duration of the study

The study was conducted in Arba Minch town, which is located about 495 km southwest of Addis Ababa, the capital city of Ethiopia. Based on the 2007 Ethiopian census report, the projected population of Arba Minch town in the year 2019/20 was 118,040 of which 50.2% were females. In the town, there are eleven kebeles (the smallest administrative unit in Ethiopia) and two youth club centers. About 5,733 youths were school enrolled of which 51.6% were male. The data were collected from August 16^th^– 30^th^, 2020.

### Study design and population

A community-based cross-sectional study was conducted among male youths in selected kebeles in Arba Minch town.

### Eligibility criteria

All male youths who lived for greater than 6 months in the town were included in the study. Those male youths who were critically ill, mentally incapacitated, and who were not able to respond to the interview questions for any reason were excluded from the study.

### Sample size sand sampling procedure

A single population proportion formula ((Zα/2) pq/d2) was used to estimate the sample size required for the study. The sample size calculation assumed the proportion (p), the estimated prevalence of attitude towards gender inequitable norms among male youths was estimated to be 50% because there is no prior study finding in Ethiopia, 95% confidence level, and margin of error of 5% which gave the sample size of 384. In consideration of a 10% non-response rate, the final sample size was 423 youths. The town has eleven Kebele administrations from these three kebeles were selected randomly by lottery method. The sample was proportionally allocated to each Kebele based on the size of the youths. Finally, the study participants were selected by simple random sampling method using data from the census as a sampling frame.

### Data collection tools and procedures

A structured questionnaire was used to collect information from study participants. The questionnaire contains six parts: socio-demographic characteristics, childhood experiences, substance use, knowledge, media use, and attitude towards gender inequitable norms. Attitude towards gender inequitable norms was measured using GEM 24 questions that were validated and translated for Ethiopia by validation study [11]. The questions for socio-demographic and childhood experience were adapted from previously published relevant works of literature [12, 13]. Data were collected by ten data collectors who were experienced and regular data collectors for the health and demographic surveillance site coordination office at Arba Minch.

### Definitions and measurements

The favorable attitude was measured using the GEM scale. For each participant, responses to the GEM scales were added together to form composite discrete variables that were categorized into high inequity = 24-39, Moderate inequity = 40-55, and low inequity = 56-72. Participants responded: Totally Agree =1; Partially Agree =2, or Do Not Agree=3 for each of the items of GEM scale Score. A mean score was used as the cut-off point to categorize the respondent as a favorable and unfavorable attitude towards gender inequitable norms. Respondents who score at the mean and below the mean were categorized as a favorable attitude towards gender inequitable norms otherwise unfavorable [9].

#### Knowledge towards gender-equitable norms

In this study, knowledge towards gender-equitable norms was measured based on the responses of the participants to six knowledge related questions that are specific to this topic. First, negatively stated questions were reverse coded during data processing. Then, it was summed up and a score above the mean is said to have good knowledge of gender-equitable norms.

#### Decision-making power

Comprises of six questions that were used to construct a composite score. Each question has three response options on the degree of women’s involvement in decision-making (mother only, the father only, and joint) in the home during childhood experience. Based on the responses, each question was scored as follows: Father only decides score = 0, Joint decision-making score =1, and women (mother) only decides score =2. After computing altogether, a score above the mean is said to have mother good decision-making power

### Data quality management

Data collectors and supervisors were provided with a daylong intensive training on the techniques of data collection and components of the instrument. Before the commencement of the data collection, a pretest was conducted among male youths (5% of the sample size) who were not included in the study. Based on the findings from the pre-test, ambiguous questions were corrected. An ongoing formative checkup for completeness and consistency of responses was made by the supervisors on a daily basis.

### Data processing and analysis

Data were entered into Epi-data software version 4.4.2.1 and then exported to SPSS version 21 statistical package for analysis. Descriptive statistics were computed and summarized in tables, figures, and text with frequencies, mean, or standard deviations where appropriate. The association between the outcome and its independent variables was examined by binary logistic regression. Variables that showed statistically significant during bi-variable analysis at p-value <0.25 were entered into a multivariable logistic regression to identify statistically significant variables. Adjusted odds ratio with 95% CI, carried out to assess the strength of the associations, and statistical significance was declared at a p-value <0.05.

### Ethics consideration and consent to participate

Ethical approval and clearance were obtained from Arba Minch University Institutional research Ethics Review Board, College of Medicine and Health Sciences. A letter of cooperation was obtained from the school of Public health. The purpose of the study was explained and informed written consent was taken from each youth and confirmation that written informed consent was obtained from the parent/guardian of each participant under 18 years of age. To ensure confidentiality, their names, and other personal identifiers were not registered in the survey tool. Besides, this study was conducted in accordance with the Declaration of Helsinki, and all ethical and professional considerations were followed throughout the study to keep participants’ data strictly confidential.

## Results

### Socio-demographic & behavioral characteristics

The data were collected from 423 study participants with a 100% response rate. The mean age of respondents was 20.24 (±S.D 2.81) years in which more than half (n=248, 58.5%) of respondents were in the age group of 20-25 yrs. Of the total study subjects, 362 (85.6%) were single. The majority, 417(93.9%) of the participants attended formal education. From the total study subjects, 117(27.6%) of the participant had 2001-4000 ETB average family income per month. Three hundred fifteen (74.5%) of participants were students (Table 1).

**Table1.** Socio-demographic & behavioral characteristics of study participants at Arba Minch town, 2020 (N=423)

| Respondents Characteristics | Category | Frequency | Percent (%) |
| --- | --- | --- | --- |
| Age | 15-19 | 175 | 41.4 |
|  | 20-25 | 248 | 58.6 |
|  | Single | 362 | 85.6 |
| Marital status | Married | 51 | 12.0 |
|  | In relationship | 10 | 2.4 |
| Educational status | No formal education | 26 | 6.1 |
|  | Primary | 85 | 20.1 |
|  | Secondary | 143 | 33.8 |
|  | Above Secondary | 169 | 40.0 |
| Mother educational status | No formal education | 184 | 43.5 |
|  | Primary | 132 | 31.2 |
|  | Secondary | 72 | 17.0 |
|  | Above Secondary | 35 | 8.3 |
| **Father educational status** | No formal education | 102 | 24.1 |
|  | Primary | 142 | 33.6 |
|  | Secondary | 89 | 21.0 |
|  | Above Secondary | 90 | 21.3 |
| **Family income** | <2000 | 99 | 23.4 |
|  | 2001-4000 | 116 | 27.6 |
|  | 4001-6000 | 105 | 24.7 |
|  | >6000 | 103 | 24.3 |
| **Occupational status** | Student |  |  |
|  | Government employed | 315 | 74.5 |
|  | Daily labor | 25 | 5.9 |
|  |  | 83 | 19.6 |

### Childhood Experience of the youth and decision making power of the mother

Around three-quarter (72.1%) of respondents were grown with both their mother and father as guardian. The remaining 14.9%, 9.5%, and 3.5% were grown with only mother, with other family members, and only father respectively. Three hundred forty-five (81.6%) of the respondents reported that they had experienced at least one form of physical violence during their childhood. Among the participants, 73.8%, 45.4%, 51.8%, 43.7%, 54.0%, and 36.4% of them were slapped, pushed, hit, kicked, choked or burnt, and stabbed with a gun or knife respectively. Two hundred five (48.5%) of youths perceived that their mother’s had good decision making power in their family.

### Substance use

Out of the study participants, 136(32.2%) of youths reported ever use alcohol, and 37.8% were currently alcohol users. Among participants, 30.5% of youths ever chew chat, and 22.2% chewing chat currently. Furthermore, 14.4% of the participant ever smokes cigarette, and 8.5 % currently smoke cigarette.

### Media usage

More than half of men participants 315 (74.5%) had information about gender equity on media. From those 33.8%, 46.4%, 9.9%, and 9.9% of participants obtain information from school, TV, Radio, and social media respectively. Furthermore, 108 (25.5%) of youths were participating in youth clubs.

### Knowledge of study participants

Two hundred fifty-five (60.3%) of the participants had poor knowledge towards gender-equitable norms (Table 2).

**Table 2:** Knowledge of study participants towards Gender inequitable norms at Arba Minch town, Ethiopia, 2020 (N = 423)

| Variables used to measure knowledge |  | Frequency (n) | Percent% |
| --- | --- | --- | --- |
| **Knew that** |  |  |  |
| All women are equal to men | Yes | 254 | 60.0 |
|  | No | 169 | 40.0 |
| “Women’s right” is a loss to men? | Yes | 66 | 15.6 |
|  | No | 357 | 84.4 |
| Woman participate in a top political position | Yes | 366 | 86.5 |
|  | No | 57 | 13.5 |
| Making all decision about the family is men’s roll | Yes | 162 | 38.3 |
|  | No | 261 | 61.7 |
| The indoor activity is only women's responsibility | Yes | 154 | 36.4 |
|  | No | 269 | 63.6 |
| Formal education is for men's only | Yes | 20 | 4.7 |
|  | No | 403 | 95.3 |
| Overall knowledge of the participants | Good | 168 | 39.7 |
|  | Poor | 255 | 60.3 |

### Attitude towards gender inequitable norms

We measured youth’s attitudes towards equitable gender norms using the Gender Equitable Men (GEM) scale which consisted of 24 items. Results revealed that youths scored a mean score of 54.4. The mean score was used to categorize the attitude as favorable and unfavorable attitude. Accordingly, from all respondents, 200(47.3%) with 95%CI (42% - 52%) had a favorable attitude towards gender inequitable norms and the remaining 223 (52.7%) of youths had an unfavorable attitude (Table 3).

**Table 3:** Items used to measure attitude towards Gender Equitable Norms Scale at Arba Minch town Ethiopia, 2020 (N=423)

| **GEM (Gender Equitable Men)** | **Totally Agree**<br>**N (%)** | **Partially agree**<br>**N (%)** | **Disagree**<br>**N (%)** |
| --- | --- | --- | --- |
| **Violence** |  |  |  |
| There are times when a woman deserves to be beaten | 211(49.9) | 102 (24.1) | 110 (26.0) |
| A woman should tolerate violence in order to keep her family together | 243(57.5) | 42 (9.9) | 138 (32.6) |
| It is okay for a man to hit his wife if she won't have sex with him | 87 (20.6) | 48 (11.3) | 288 (68.1) |
| It is alright for a man to beat his wife if she is unfaithful | 145 (34.3) | 105 (26.0) | 168 (39.7) |
| If someone insults a man he should defend his reputation with force | 109 (25.8) | 55 (13.0) | 259 (61.2) |
| A man using violence against his wife |  |  |  |
| is a private matter that shouldn't be discussed outside | 180 (42.6) | 75 (17.7) | 168 (39.7) |
| **Domestic Life and Childcare** |  |  |  |
| A man should have the final word on decisions in his home | 227 (53.7) | 90 (21.1) | 106 (25.2) |
| A woman's most important role is to take care of her home and cook for her family | 194(45.9) | 86 (20.3) | 143 (33.8) |
| Giving the kids a bath and feeding the kids are only the mother's responsibility | 201(47.5) | 86 (20.3) | 136 (32.2) |
| A woman should obey her husband in all things | 248 (58.6) | 81 (19.2) | 94 (22.2) |
| A man should not take his child to the clinic without the child's mother | 39 (9.2) | 35 (8.3) | 349 (82.5) |
| **Reproductive Health and Disease Prevention** |  |  |  |
| It is a woman's responsibility to avoid getting pregnant | 99 (23.4) | 86 (20.3) | 238 (56.3) |
| A real man produces a male child | 357 (84.4) | 28 (6.6) | 38 (9.0) |
| Only when a woman has a child is she a real woman | 36 (8.5) | 29 (6.9) | 358 (84.6) |
| Women who carry condoms on them are easy | 113 (26.7) | 50 (11.8) | 260 (61.5) |
| A man should be outraged if his wife asks him to use a condom | 50 (11.8) | 39 (9.2) | 334 (79.0) |
| **Sexuality** |  |  |  |
| Men need more sex than women do | 210 (49.6) | 77 (18.2) | 136 (32.2) |
| You don't talk about sex, you just do it | 71 (16.8) | 46 (10.9) | 306 (72.3) |
| Men are always ready to have sex | 116 (27.4) | 81 (19.2) | 226 (53.4) |
| Real men do not immediately go a doctor when they are sick | 64 (15.1) | 40 (9.5) | 319 (75.4) |
| It is the man who decides when to have sex with a partner | 100 (23.6) | 55 (13.0) | 268 (63.4) |
| Men need other women even if everything with his wife is fine | 33 (7.8) | 27 (6.4) | 363 (85.8) |
| Employed women do not make a good wife | 69 (16.3) | 32 (7.6) | 322 (76.1) |
| A woman should not initiate sex | 104 (24.6) | 78 (18.4) | 241 (57.0) |

### Factors associated with the attitude towards gender inequitable norms

Multivariable logistic regression analysis was performed and five variables namely: educational level, household income, participation in youth clubs, knowledge towards gender inequitable norms, and physical violence during childhood were independently associated with a favorable attitude towards gender inequitable norms. Male youths with no formal education were 5.4 times [AOR= 5.4 (95% CI: 1.53, 16.5)] more likely to have a favorable attitude towards gender inequitable norms compared with a male with the educational level of above secondary. Male youths in the primary school were 2.3 times [AOR=2.3, (95%CI 1.05, 5.29)] more likely to have a favorable attitude towards gender inequitable norms compared with male youths with the educational level of above secondary. Household income less than or equal to 2,000 birr were 6 times [AOR= 6.07, (95% CI 2.91, 12.72)] more likely to have a favorable attitude towards gender inequitable norms compared with household income 6,000 and above birr. Household income with a range of 2,001 – 4,000birr were 4.2 times [AOR= 4.2; (95% CI, 2.08, 8.67)] more likely to have a favorable attitude towards gender inequitable norms compared with household income 6,000 ETB and above. Likewise, Male youths who had experienced physical violence during childhood [AOR= 2.8; (95% CI, 1.45, 5.445)], having poor knowledge [AOR=4.07; (95% CI, 2.451, 6.77)], and male youths who didn’t participate in youth club [AOR=3.6; (95% CI, 1.98, 6.68)] were more likely to have a favorable attitude towards gender inequitable norms compared with their counterparts (Table 4).

**Table 4:**
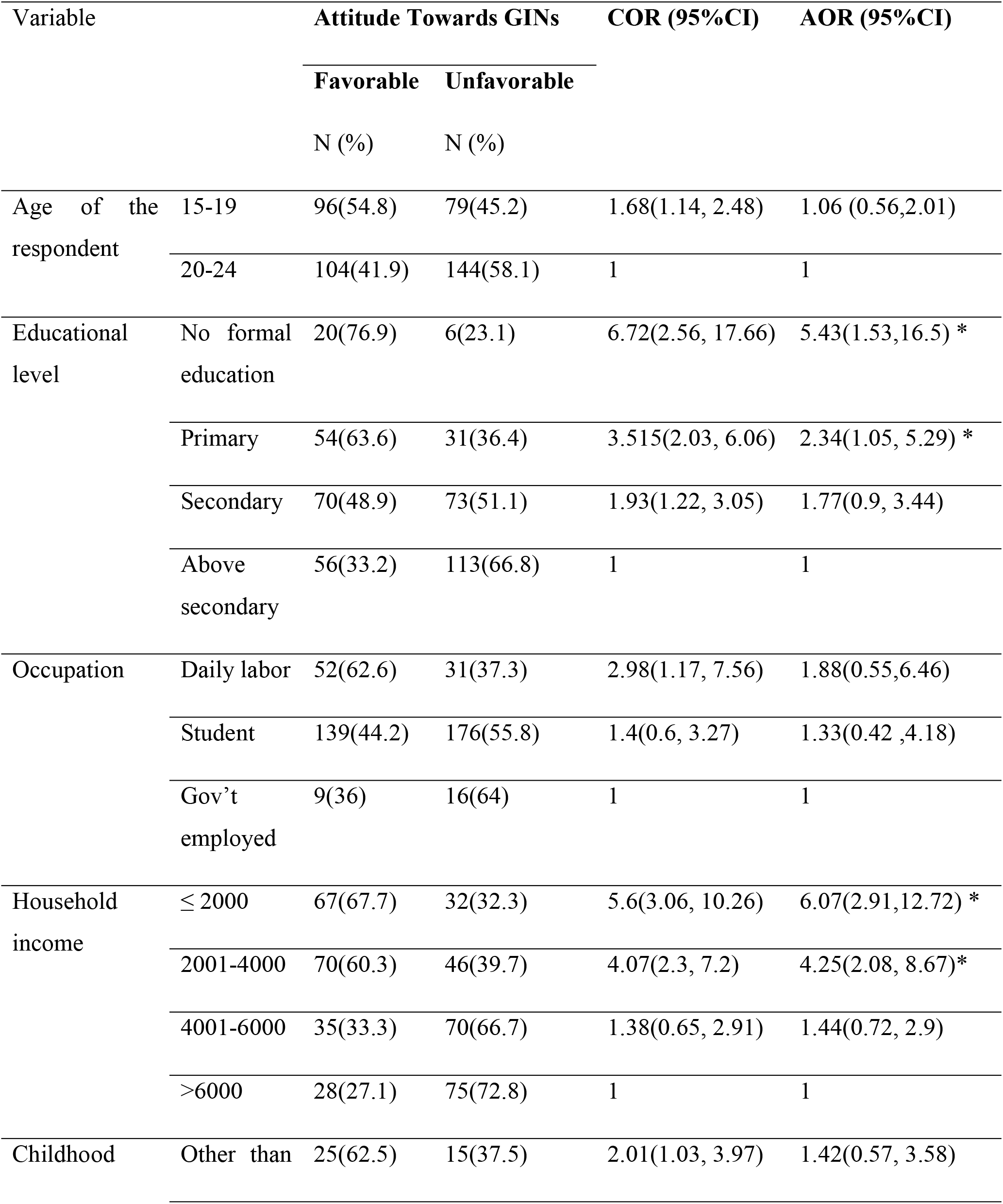
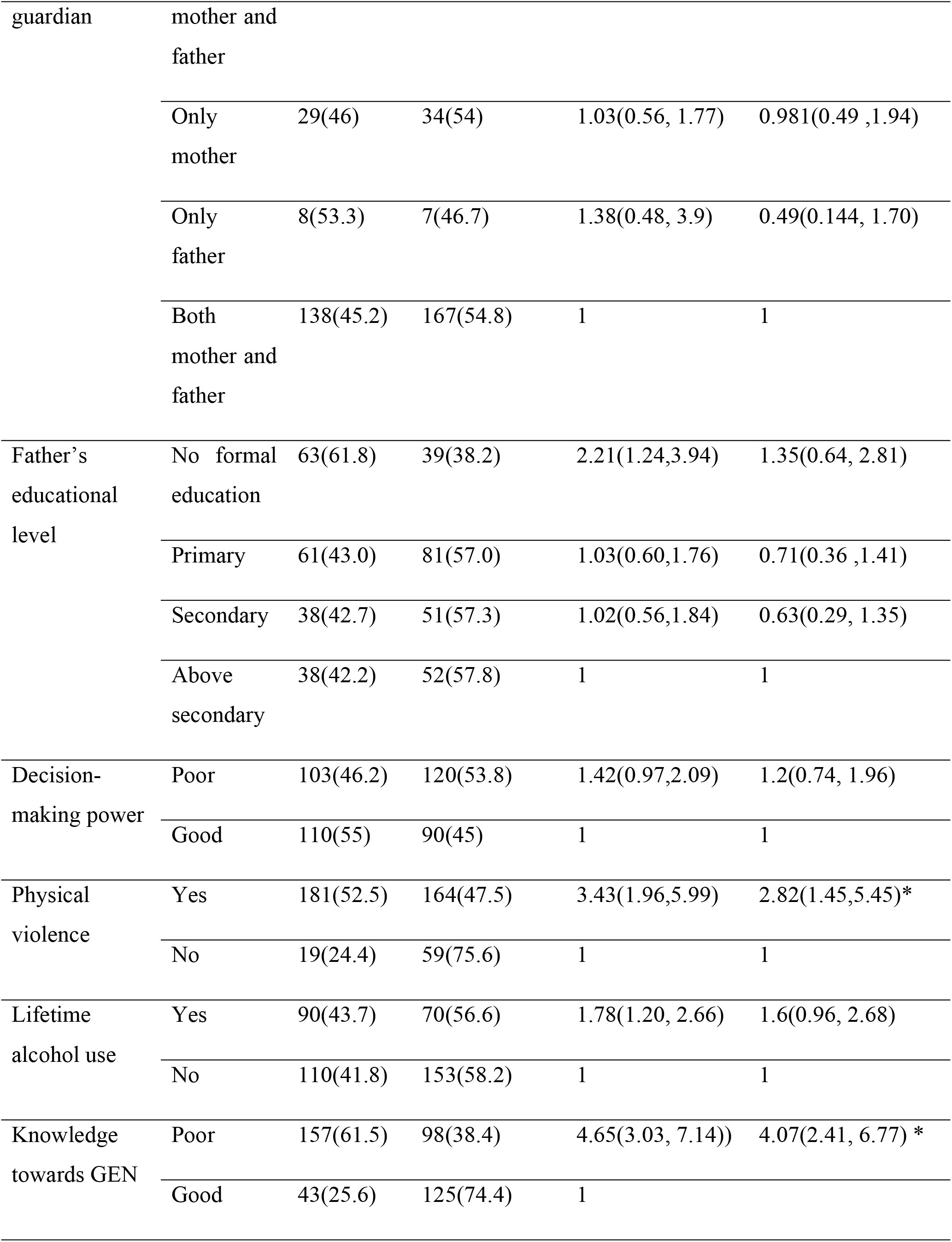
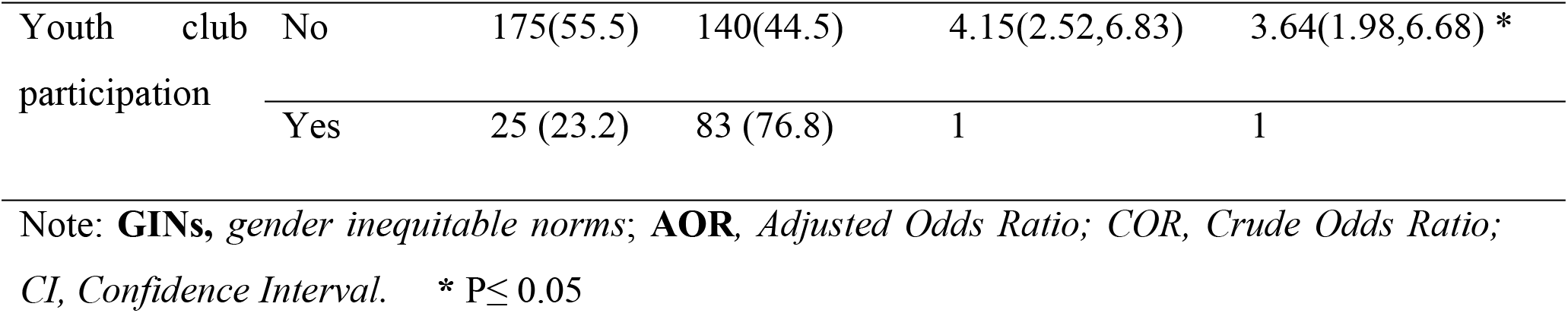
Bi-variable and Multi-variable binary logistic regression analysis for factors associated with Attitude towards gender inequitable norms at Arba Minch town Ethiopia, 2020(N=423)

## Discussion

This research is aimed to assess attitudes towards gender inequitable norms and associated factors among male youths in Arba Minch town Southern Ethiopia. In this study, the favorable attitude towards GINs is 47.3% with 95%CI (42% - 52%). It is lower than a report from a study conducted in Uganda and Ethiopia that reported favorable attitudes towards gender inequitable norms were 63.3% and 59.9% respectively. The variations might be due to the difference in the study areas, the assessment period, the tools and methods used for the studies [14, 15]. However, the favorable attitude of this study is higher when compared with studies conducted in Congo (38.7%) [9]. The variation might be due to differences in cultural characteristics of the study population. In the Congo study, the population for the sample was taken from a church going men.

More than 67% of youth participants had agreed on bathing and feeding kids are only the mother’s responsibility. This finding is lower when compared with the studies conducted in Uganda, which was 84% [16]. From all participants, 91% of youths had agreed on “a real man produces a male child” on GEMS. This finding is consistent with a study conducted in Nepal and Vietnam [17]. This may be because both countries’ cultures consider the boy as a shield to his house and the need to carry on the family name. Besides, most of the local traditional proverbs in the community also support son preference attitude.

Male youths with no schooling were more likely to have a favorable attitude towards GINs compared with the male youth with above secondary educational level. Likewise, males with the primary school were more likely to have a favorable attitude towards gender inequitable norms compared with those with above secondary educational level. This finding is consistent with the study conducted in Jamaica and Brazil [13, 18]. This may be because as youth’s educational level increases access to information; their awareness and struggle against the traditional gender norm will increase and also improve their interpersonal skill and healthier communicative relationship.

Furthermore, the participant’s average income of family has also an association with the attitude towards gender inequitable norms. It was significantly higher among households with low economic status compared with those who had high economic status. This was in line with other studies conducted in Botswana Swaziland [19], which revealed low economic status had a significant association with a favorable attitude towards gender inequitable norms. This may be because the majority of the household who had low income cannot meet their daily needs and afford to pay for school, so they engage in daily labor and remain uneducated. Resources are important for sustained behavioral change and lack of resources might be a limitation for a lifestyle change.

Males who have experienced physical violence during their childhood were three times more likely to have a favorable attitude towards GINs compared with their counterparts. This finding is supported by the studies conducted in the Democratic Republic of Congo [20]. This might be due to negative childhood experience have a significant impact on the construction of negative gender perception when they grew up.

Knowledge of males about gender-equitable norms was statistically associated with a favorable attitude towards gender inequitable norms. As male youth knowledge increase on gender inequitable norms, the odds of favorable attitude towards gender inequitable norms decrease. This study revealed that males who had poor knowledge towards gender-equitable norms were four times more likely to have a favorable attitude towards gender inequitable norms compared with those males who had good knowledge towards gender equity. Even though there is a limitation of evidence that shows an association between knowledge towards gender inequitable norms and attitudes towards gender inequitable norms, a study conducted in Nepal and Vietnam on gender, masculinity, and son preference revealed men’s knowledge towards policies and laws promoting gender equity has a significant correlation to attitudes on gender equity. The reason might be having knowledge about gender equity is a prerequisite to a favorable attitude towards gender-equitable norms. The shreds of evidence supported by a study on male youth norms initiative in Ethiopia revealed that improving awareness in men youth’s successfully influenced participants’ attitudes towards gender norms [14].

Male who did not participate in youth clubs were more likely to have a favorable attitude towards gender inequitable norms compared with those who did participate in the youth clubs. In this study youth club participation has revealed a significant association with a favorable attitude towards gender inequitable norms. Some of the possible reasons could be those who participated in the youth club might have a chance to engage in discussions or a variety of training such as the negative impact of early marriage, gender-based violence, and other gender equity-related topics. These types of exposure or training might have a factor for them to have a more favorable attitude towards gender-equitable norms.

## Limitation of the study

The following limitations should be considered when interpreting the findings; some sensitive questions like sexual domain in GEMS and substance use might introduce social desirability bias. So, to mitigate these limitations, privacy was maintained by interviewing them alone and the data collectors were men to minimize the social desirability bias. On the other hand, there could be a recall bias because men were asked to recall their childhood experience on physical violence, women’s autonomy, and others.

## Conclusions and Recommendations

The findings of this study showed that almost half (47.3%) of male youth had a favorable attitude towards gender inequitable norms. Low level of education, childhood physical violence, and poor knowledge towards gender-equitable norms, low-income household, and not participating in youth club was found to be statistically significant factors associated with a favorable attitude towards gender inequitable norms. Proactive measures are needed to curb the problem by taking integrated interventions for youths like enrolling in school, enhancing participation in a youth club, prevent childhood physical violence, increasing household income, and creating awareness on gender inequitable norms to change traditional beliefs that undermine women.

## Data Availability

The data used and/or analyzed during the current study available from the corresponding author on reasonable request.

## Acknowledgments

The authors are thankful for the data collectors, the study participants, and supervisors for their cooperation during data collection. Our thanks also go to Arba Minch University for providing ethical clearance.

## Authors Contributions

**Conceptualization:** Barkot Samuel, Muluken Basa, Mulugeta Shegaz, and Zeleke Gebru

**Data curation:** Barkot Samuel, Muluken Basa, Mulugeta Shegaz, Zeleke Gebru, Mustefa Glagn

**Formal analysis:** Barkot Samuel, Muluken Basa, Mulugeta Shegaz, and Zeleke Gebru

**Funding acquisition:** Barkot Samuel, Mulugeta Shegaz, Zeleke Gebru and Belay Boda

**Investigation:** Barkot Samuel, Mulugeta Shegaz, Muluken Basa and Zeleke Gebru

**Methodology:** Barkot Samuel, Mulugeta Shegaz, Muluken Basa, Zeleke Gebru, and Mustefa Glagn

**Project administration:** Barkot Samuel, Mulugeta Shegaz, Muluken Basa, Zeleke Gebru.

**Resources:** Barkot Samuel, Mulugeta Shegaz, Muluken Basa, Zeleke Gebru and Mustefa Glagn

**Software:** Barkot Samuel, Mulugeta Shegaz, Muluken Basa, Zeleke Gebru, Belay Boda

**Supervision:** Barkot Samuel, Mulugeta Shegaz, Muluken Basa, Zeleke Gebru and Belay Boda

**Validation:** Barkot Samuel, Mulugeta Shegaz, Muluken Basa, Zeleke Gebru

**Visualization:** Barkot Samuel, Mulugeta Shegaz, Zeleke Gebru, Belay Boda and Mustefa Glagn

**Writing – original draft:** Barkot Samuel, Mulugeta Shegaz, Muluken Basa, Zeleke Gebru, and Mustefa Glagn

**Writing – review & editing:** Barkot Samuel, Mulugeta Shegaz, Muluken Basa, Zeleke Gebru, and Mustefa Glagn

## Data Availability

The datasets used and/or analyzed during the current study available from the corresponding author on reasonable request.

